# Geographical and temporal variation in reduction of malaria infection among children under five years of age throughout Nigeria

**DOI:** 10.1101/2021.01.06.21249363

**Authors:** Wellington Oyibo, Godwin Ntadom, Perpetua Uhomoibhi, Olusola Oresanya, Nnenna Ogbulafor, Olufemi Ajumobi, Festus Okoh, Kolawole Maxwell, Sonachi Ezeiru, Ernest Nwokolo, Chioma Amajoh, Nnenna Ezeigwe, Mohammed Audu, David J. Conway

## Abstract

**Background:** Global progress in reducing malaria has stalled since 2015. Analysis of the situation is particularly needed in Nigeria, the country with by far the largest share of the burden, where approximately a quarter of all cases in the world are estimated to occur.

**Methods:** We analysed data from three nationwide surveys (Malaria Indicator Surveys in 2010 and 2015, and a National Demographic and Health Survey in 2018), with malaria parasite prevalence in children under five years of age determined by sampling from all 36 states of Nigeria, and blood slide microscopy performed in the same accredited laboratory for all samples. Changes over time were evaluated by calculating prevalence ratio (PR) values with 95% confidence intervals (CI) for each state, together with Mantel-Haenszel adjusted prevalence ratios (PR_adj_) for each of the six major geopolitical zones of the country.

**Results:** Between 2010 and 2018 there were significant reductions in parasite prevalence in 25 states, but not in the remaining 11 states. Prevalence decreased most in southern zones of the country (South West PR_adj_ = 0.53; South East PR_adj_ = 0.59; South South PR_adj_ = 0.51) and the North Central zone (PR_adj_ = 0.36). Changes in the north were less marked, but were significant and indicated overall reductions by more than 20% (North West PR_adj_ = 0.74; North East PR_adj_ = 0.76). Changes in the south occurred mostly between 2010 and 2015, whereas those in the north were more gradual and most continued after 2015. Recent changes were not correlated with survey-reported variation in use of preventive measures.

**Conclusion:** Reductions in malaria infection in children under five have occurred in most individual states in Nigeria since 2010, but substantial geographical variation in the timing and extent indicate challenges to be overcome to enable global malaria reduction.

## Introduction

Currently, half of the global malaria burden is caused by Plasmodium falciparum in West Africa, most substantially in Nigeria which is estimated to have approximately a quarter of all cases in the world and a similar proportion of the overall malaria mortality ^1 2^. Under a ‘High burden to high impact’ initiative, the World Health Organization (WHO) has issued a ‘wake-up call’ noting the need for urgent targeted action ^3^, particularly relevant to Nigeria where WHO-estimated numbers of cases are increasing rather than reducing annually ^2 3^ Knowing how to reduce malaria in the future clearly requires understanding of current epidemiology, and identification of changes that have already occurred ^4^.

Notable reductions in malaria burden have been seen in some areas of West Africa, especially near the north-western edge of the endemic distribution in Senegal and The Gambia ^5-7^, encouraging studies to evaluate whether local elimination might become feasible in future ^8^. However, there is an even more pressing need to understand why the malaria burden remains high in other parts of the region, particularly in Nigeria. This is a priority to reduce global mortality and morbidity, and to reduce the main reservoir which may prevent other countries from achieving elimination due to continued importation of infections ^9^.

Malaria surveillance at community level is important, as this has the potential to give unbiased indication of geographical variation and trends over time, but such data are lacking from most areas so that trends are only estimated by modelling ^10^. Separate small-scale surveys conducted in some communities in Nigeria over the past decade have indicated continuing high levels of malaria parasite infection (almost all P. falciparum), particularly in areas of the north with prevalence still exceeding 60% ^11-13^, almost as high as reported in similar areas half a century or longer ago ^14 15^. In contrast, surveys in some areas in the south of the country report much lower infection prevalence, although with considerable local variation as illustrated by surveys in different communities within Lagos state ^16-18^. National Malaria Indicator Survey (MIS) data from 2010 and 2015 included slide microscopy data from children under five years of age, and these also showed regional variation and confirmed that states in the north of the country generally had higher prevalence than in the south ^19 20^. The 2015 data showed lower prevalence overall than was seen in the 2010 survey ^20^, although analysis within the earlier survey report was limited to broad geographical zones rather than for each of the individual states sampled.

Recent availability of results from a National Demographic and Health Survey (NDHS) based on sampling from all 36 Nigerian states in 2018 ^21^, together with the previous MIS data from 2010 and 2015, means that for the first time there are data for Nigeria from three nationwide surveys in which malaria parasite slide prevalence in children under the age of five years has been measured. There are two important features of each of these three national surveys, for the purpose of critically analysing trends in infection. Firstly, they were conducted at a similar period in each year, which minimises confounding of comparisons due to seasonal variation: 2010 MIS survey (October to December) ^20^, 2015 MIS Survey (October to November) ^19^, 2018 NDHS survey (August to December) ^21^. Secondly, but equally importantly, slide microscopy for each of these surveys was conducted under quality-controlled conditions in the same accredited laboratory.

Here, we analysed the microscopy data from these three nationwide surveys, and evaluated changes over time by calculating prevalence ratios between the surveys for each of the different states, together with adjusted prevalence ratios for each of the major geographical zones of the country. Emerging from this analysis are details of trends in reduction of malaria prevalence that are informative for formulating future intervention strategies. This also highlights opportunities for additional measurements to be made as part of future nationwide surveys, to help identify means of effectively reducing the large malaria burden.

## Methods

### Population under analysis

The population of Nigeria was estimated by the Nigerian Population Commission to have exceeded 182 million by late 2016, and is currently estimated by the UN as over 200 million, within a land area of 923,768 km^2^. Malaria occurs throughout the country in highly diverse ecological zones existing in succession from south to north: Mangrove Swamp and Coastal Vegetation, Freshwater Swamp Forest, Lowland Rain Forest, Derived Savanna, Guinea Savannah, Sudan Savanna, and Sahel Savanna. The country is divided into 36 states (Figure 1) grouped into six geopolitical zones: North -East, North-Central, North-West, South-East, South-South and South-West, each of which contains between five and seven states (Table 1), apart from the Federal Capital Territory (FCT) of Abuja. The National Malaria Elimination Programme of the Federal Ministry of Health provides policy and guidance for malaria control in Nigeria, while the implementation of interventions is done at the individual state level. The rate of implementation differs from one state to the other, influenced by factors including political will, resource mobilization, and partnership support, which are not easily quantifiable, although population-based surveys can provide some indices of reported coverage and use of implementations.

**Table 1.**
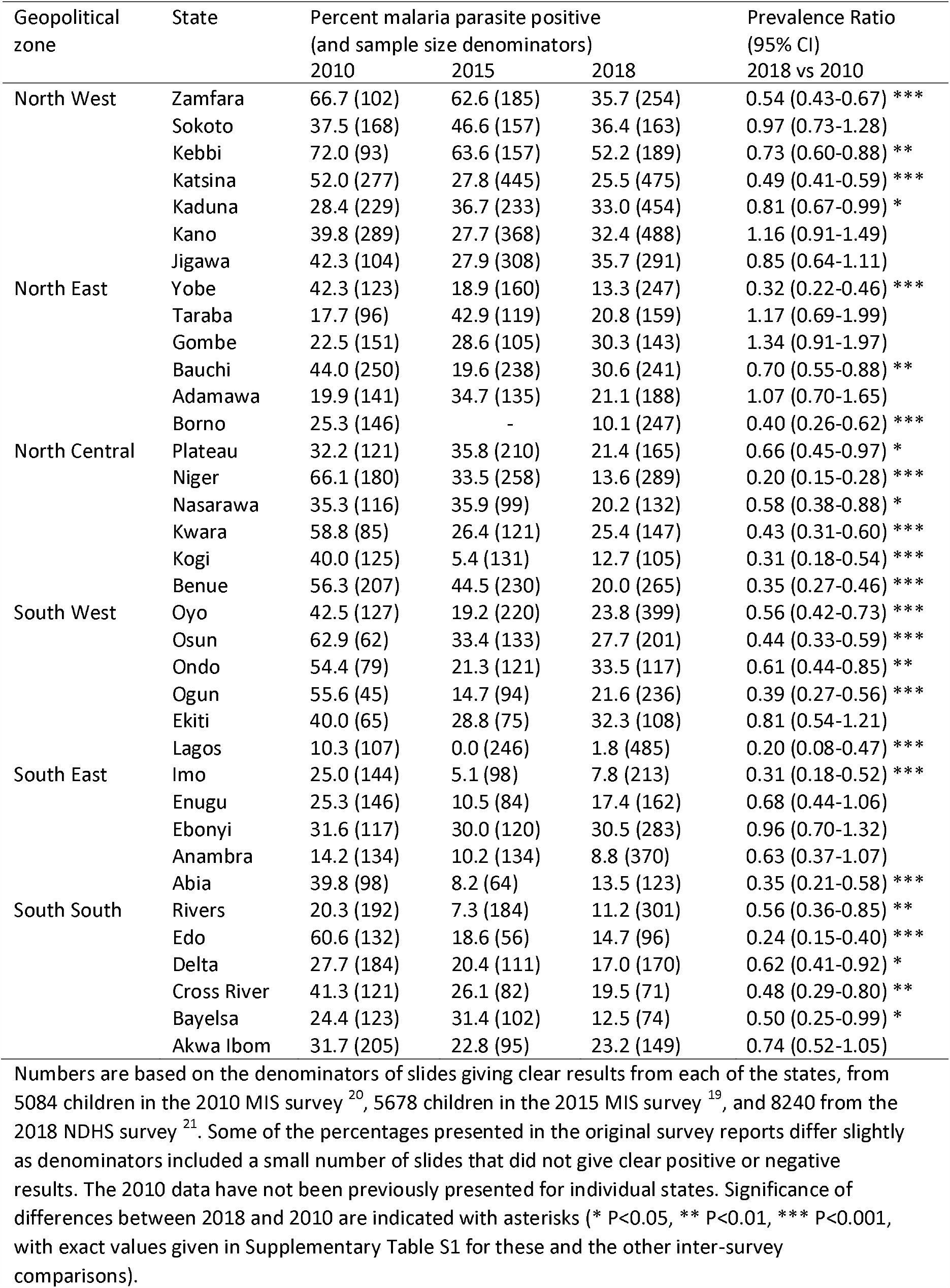
Malaria slide positivity in children under 5 years of age in three nationwide community surveys in Nigeria using standardised quality-controlled microscopy procedures

**Figure 1.**
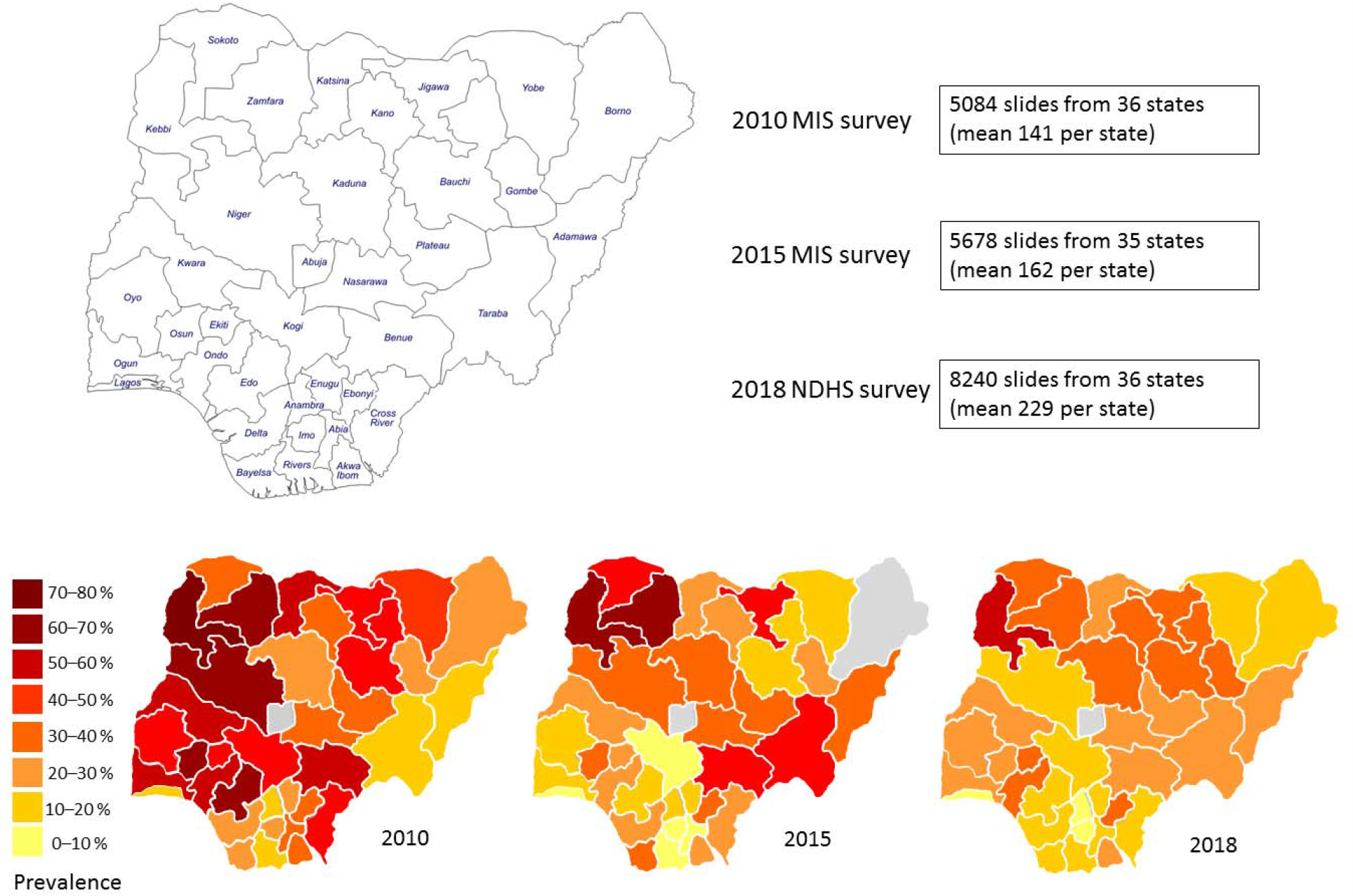
Geographical and temporal heterogeneity of malaria parasite prevalence in children under five years old in Nigeria. **A**. Data are analysed for all 36 states (excluding Borno state in 2015); the central Federal Capital Territory incorporating Abuja is not analysed as it could not be representatively sampled. **B**. The names of individual states are shown, and the grouping into six geopolitical zones is given in Table 1. **C**. Data are derived from three previous surveys, made available for analysis from http://mics.unicef.org/surveys with permission by the UNICEF MICS team. All numbers are given in Table 1 and Supplementary Table S1.

### Data from nationwide population-based surveys

Three nationwide population-based surveys of malaria infection have been conducted in Nigeria, as part of two Malaria Indicator Surveys (MIS) and a recently published National Health and Demographic Survey (NDHS). Standardised methods of household cluster sampling were performed within the 2010 MIS survey (October to December) ^20^, 2015 MIS Survey (October to November) ^19^, and 2018 NDHS survey (August to December) ^21^. In the 2015 and 2018 surveys random cluster sampling methodology was performed to select clusters of households within each of the separate states. In the 2010 survey, randomisation was performed within each of the six major geographical zones of the country, but there was a wide dispersal of sampled clusters within each of the 36 separate states and overall sample sizes were only slightly lower than in the later surveys, leading to no substantial bias and broadly similar accuracy for the purpose of the current retrospective analysis. The protocols for the surveys were approved by the National Health Research Ethics Committee of Nigeria and the Ethics Committee of ICF, USA. All data were generated under the Demographic and Health Surveys (DHS) Program (https://dhsprogram.com/), and have been made available (http://mics.unicef.org/surveys) with permission granted by the UNICEF MICS team.

### Malaria parasite microscopy

In all surveys, finger prick samples of capillary blood from children under 5 years of age (6-59 months) were taken to prepare thick and thin peripheral blood films for microscopy, using barcoded slides that were dried and stored in labelled slide boxes, and transported to the ANDI (African Network for Drugs and Diagnostics Network) Centre of Excellence for Malaria Diagnosis, College of Medicine, University of Lagos (a well-recognised accredited reference centre for multicentre diagnostic research with microscopy gold standard measurement ^22^). Upon reception, the barcoded slides were individually scanned for reading into an electronic database created for the survey, and stained with 3% Giemsa using the WHO-recommended standard operating procedure MM-SOP-04. Microscopical slide reading according to the protocol recommended by WHO for detection of malaria parasites was done independently by two WHO-Certified Malaria Grade Level 1 Microscopists. To monitor the independence of the slide reading, the process was managed by a Slide Coordinator who received the microscopy results from each microscopist, and reviewed results for concordance. Where there was discordance in slide positivity, a third Certified Grade Level 1 Microscopist performed an independent slide read to provide resolution. The malaria microscopy data were entered independently by two data entry clerks to the Census Survey Processing System (CSPro software database, United States Census Bureau, USA), within the original survey protocols.

### Statistical analysis of slide microscopy data from each of the different surveys

Slide microscopy data from the ANDI laboratory generated for the three separate nationwide surveys are analysed for trends in each of the 36 states nationwide. From the 2010 MIS survey, slide microscopy results for 5084 children in the 36 states were analysed, giving a mean sample size of 141 per state. From the 2015 MIS survey, slide microscopy results for 5678 children in 35 states were analysed, a mean sample size of 162 per state (Borno state was omitted from analysis for 2015 as it was not possible to perform representative sampling due to security challenges). From the 2018 NDHS survey, slide microscopy results for 8240 children in the 36 states were analysed, giving a mean sample size of 229 per state. Changes over time were analysed by calculating prevalence ratio (PR) values with 95% confidence intervals (CI) between each of the different surveys for each of the different states, together with Mantel-Haenszel adjusted prevalence ratios (PR_adj_) and CI each of the six major geographical zones of the country. Testing for correlations between temporal prevalence ratios and reported use of insecticide treated nets (ITNs) was performed using Spearman’s non-parametric rank correlation across all states. Analyses of the data here were performed using SPSS and EPI-INFO, with graphical presentations using PRISM.

### Patient and Public Involvement

This study is a comparative analysis of three nationwide surveys conducted from 2010 to 2018 with sampling under previous protocols, and all analyses are based on metapopulation comparisons of data from 36 states, so it was not appropriate to involve patients or particular public communities in the design, conduct, reporting, or dissemination plans.

## Results

From the nationwide surveys in 2010, 2015 and 2018, blood slide microscopy results allow estimation of malaria parasite infection prevalence in children under 5 years of age. The original survey reports indicated the overall nationwide prevalence as declining from 42% in 2010 to 27% in 2015 ^19 20^, with only a modest subsequent decline to 23% in 2018 ^21^. Data were here analysed in more detail for all 36 states of Nigeria in each of the survey years (Figure 1 and Table 1), excluding Borno state in 2015 for which there was insufficient data. We checked denominators so that only individuals having slides with a valid read result were included (a few individuals without a clear slide result were previously included in some previous denominators in original survey reports but were not counted in analysis here).

The malaria parasite prevalence in children under 5 years of age in each of the surveys shows marked geographical patterns and temporal differences (Figure 1 and Table 1). Focusing on areas with high prevalence, 17 of the states had prevalence of above 40% in 2010, many in northern and western areas but also some in other areas of the country, whereas in 2018 only one state had prevalence of above 40% (Kebbi state in the north-western part of the country). Conversely, focusing on areas with less malaria, in 2010 only five states had prevalence of less than 20%, all in southern and eastern areas, whereas by 2018 there were 14 states with prevalence of less than 20% (Figure 1 and Table 1).

Statistical analysis of the data from each of the surveys uncovers more details on the magnitude and timing of the changes, and enable these to be evaluated not only for the major geopolitical zones of the country, but also for the individual states. Between 2010 and 2018 there were statistically significant declines in prevalence in 25 out of 36 states, and no significant change in prevalence in the remaining 11 states (Table 1 and Figure 2). The declines were greatest in the three major southern zones of the country and in the North Central zone, with Mantel-Haenszel adjusted prevalence ratios (PR_adj_ with 95% confidence intervals, CI) indicating overall relative reduction by more than 40% (South West PR_adj_ = 0.53, CI 0.46 - 0.61; South East PR_adj_ = 0.59, CI 0.49 - 0.72; South South PR_adj_ = 0.51, CI 0.43 - 0.61; North Central PR_adj_ = 0.36, CI 0.32 - 0.42). Although the declines in the northern zones were less marked and not significant in many states, they were still significant overall, indicating relative reduction by more than 20% (North West PR_adj_ = 0.74, CI 0.68 - 0.81; North East PR_adj_ = 0.76, CI 0.65 - 0.89).

**Figure 2.**
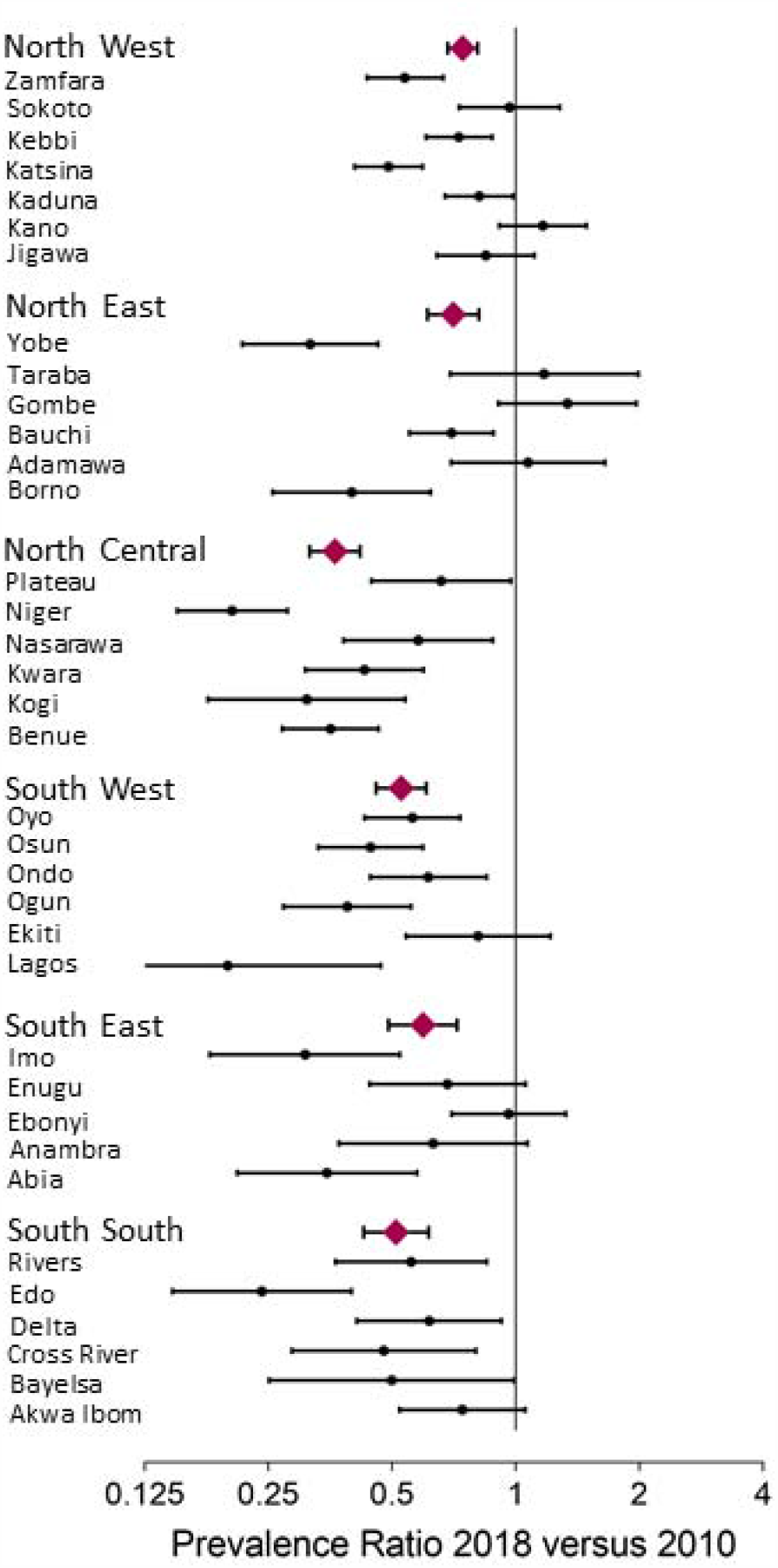
Prevalence ratios (with 95% confidence intervals) comparing malaria parasite infection in under-5 year old children in 2018 with 2010 in each state and geographical zone in Nigeria. Although there was marked variation, significant declines in prevalence were seen in many states, indicated by ratios with 95% confidence intervals to the left of the line. For each of the six major geographical zones of the country, the overall Mantel-Haenszel adjusted Prevalence Ratio (PR_adj_) is plotted with a purple diamond symbol (with 95% confidence intervals), showing significant overall declines: North West PR_adj_ = 0.74 (0.68 - 0.81); North East PR_adj_ = 0.70 (0.61 - 0.81); North Central PR_adj_ = 0.36 (0.32 - 0.42); South West PR_adj_ = 0.53 (0.46 - 0.61); South East PR_adj_ = 0.59 (0.49 - 0.72); South South PR_adj_ = 0.51 (0.43 - 0.61). All numbers and P values are given in Table 1, and individual state estimates are given in Supplementary Table S1.

Analysis of the changes between each of the successive surveys showed significant variation in the trends over time (Figure 3). Out of 35 states analysed (Borno was excluded for 2015), 20 states had statistically significantly lower prevalence in 2015 versus 2010 (Figure 3), while only 2 states had significantly higher prevalence in 2015 (Adamawa and Taraba, in the North East). The reduction in prevalence between 2010 and 2015 was significant in all of the major zones of the country, except for the North East (Figure 3). There was a greater relative reduction in the south of the country, so that the overall heterogeneity among states nationwide was increased, as reflected by the coefficient of variation (standard deviation divided by the mean, increasing from 0.411 in 2010 to 0.558 in 2015).

**Figure 3.**
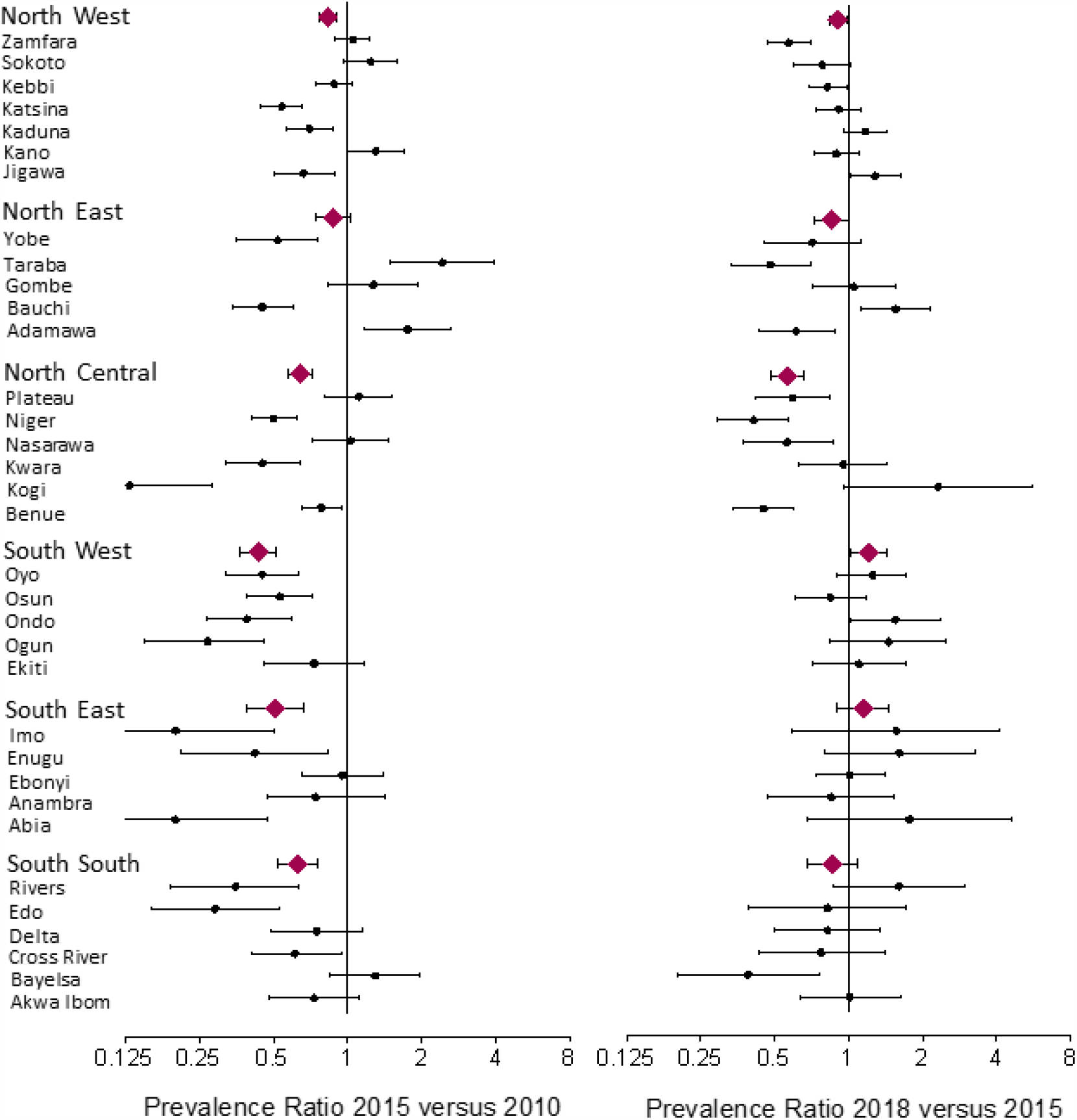
Decrease in malaria prevalence in under-5 year old children shows significant heterogeneity over time and geographical area. Prevalence ratios (with 95% confidence intervals) compare the malaria parasite infection in under-5 year old children in individual states and geographical zones of Nigeria in the intervals between 2010 and 2015 (left hand panel), and between 2015 and 2018 (right hand panel). For each of the six major geographical zones of the country, the overall Mantel-Haenszel adjusted prevalence ratio (PR_adj_) is plotted with a purple diamond symbol with 95% confidence intervals. Between 2010 and 2015 significant declines are seen in all zones, except the North East where the decline is borderline significant, and the declines are more marked in the south of the country. Between 2015 and 2018, significant declines were seen overall in North Central zone and North West zone, borderline significant in North East. (Data for Lagos state are not plotted as there was an indefinite prevalence ratio due to zero positive slides in 2015, and Borno state is not included due to lack of data for 2015).

The comparison of subsequent differences between 2015 and 2018 in these 35 states shows that 9 states had statistically significantly lower prevalence in 2018, while 4 states had significantly higher prevalence in 2018 (Figure 3 and Table 1). In contrast with the previous period, there was minimal reduction in prevalence in southern areas between 2015 and 2018, and a slight increase was seen in the South West zone (adjusted Prevalence Ratio = 1.21, 95% CI 1.01-1.43), and in some other individual states. However, there were reductions in prevalence in many of the northern states between 2015 and 2018, and significant declines were seen overall in the North Central zone (PR_adj_ = 0.56, CI 0.48 - 0.56) and North West zone (PR_adj_ = 0.90, CI 0.83 - 0.98), while the decline was of borderline significance in the North East (PR_adj_ = 0.85, CI 0.72 - 1.00). These modest but significant declines in prevalence in northern areas, where malaria burden has historically been highest, slightly reduced the overall heterogeneity across states nationwide (signified by a coefficient of variation of 0.558 in 2015 decreasing to 0.454 in 2018).

The heterogeneity in malaria trends in different parts of Nigeria should be considered in relation to changes in coverage and use of antimalarial prevention and treatment. Although it is difficult to obtain accurate data on local variation at the population level, the MIS and NDHS surveys included relevant questionnaires that at least allow trends in the household respondent data to be investigated. Nationwide, there was a substantial and continued increase in the reported proportion of malaria cases that respondents claimed were treated with a recommended artemisinin-based combination therapy (ACT), from only 6.0 % (in 2010) to 37.6 % (in 2015) and 52.0 % (in 2018) ^19-21^. As the proportions increased in parallel in all six major zones of the country, reported ACT use does not explain the observed variation in malaria trends nationwide including the recent reversals in parts of the country. Over the same period the proportions of households with respondents reporting ownership of insecticide treated nets (ITNs) increased nationwide from 42.0% (in 2010), to 69.0 % (in 2015) but then declined to 60.6 % (in 2018) ^19-21^. As ownership does not equate to use, it is notable that the reported overall level of use by children under five years of age increased from 37.3 % in 2015 to 43.2 % in 2018 ^21^, so we investigated whether variation in levels of use in each state (tabulated in Supplementary Table S1) correlated with recent changes in malaria prevalence. The prevalence ratio in 2018 compared to 2015 across individual states (as shown in Figure 3 and tabulated in Supplementary Table S1) did not correlate significantly with the reported ITN use by children under five years of age in these states in 2015 (Spearman’s rho = -0.25, P = 0.15), or in 2018 (Spearman’s rho = -0.09, P = 0.61), or with the change in ITN use reported between the surveys (Spearman’s rho = 0.26, P = 0.15).

## Discussion

Although Nigeria is the country with by far the largest burden of malaria globally ^2^, it is now clear that over the past decade there have been significant reductions in malaria infection prevalence in children under five years of age in all the major geopolitical zones of the country, and in most of the individual states. However, the relative reductions have been modest, and show substantial sub-national variation in their timing and extent. This highlights major ongoing challenges to achieve and sustain reductions in malaria infection, vital for global targets.

Considering where malaria control is most needed, it is notable that six of the states in northern Nigeria with malaria burdens that are among the highest globally had a similar prevalence in 2018 compared with 2010. It is of highest priority to understand reasons for the lack of decline in malaria infection in children under five years of age in these states, and to identify whether targeted increases in coverage of existing interventions will be effective, or whether new interventions are required. Conversely, many states in southern areas of the country had substantially reduced prevalence in 2015 compared to 2010, but no further reduction was seen by 2018, which raises concerns on how to consolidate and continue to reduce malaria after some initial progress. It was previously shown that variation in prevalence among states in the 2015 survey was greater than the sub-national variation seen in comparable DHS surveys of infection in young children other highly endemic West African countries ^23^, emphasising the need to understand divergent trends occurring within Nigeria.

It is important to highlight varying levels and timing of declines in malaria prevalence in young children, but also to acknowledge that there may be multiple causes. The NDHS survey-reported use of appropriate ACT therapy for malaria case management was higher in 2018 than in the previous MIS surveys, although the actual use may be lower than reported, and non-recommended or unofficial therapy is still very common ^21^. Although distribution and ownership of donated ITNs increased in most areas between 2010 and 2015, there was a slight decline in reported ownership of ITNs by 2018, and levels of use remain low in many communities throughout the country. Although there was no significant correlation between reported ITN use in children under five and the variable trends in malaria infection in this age group between 2015 and 2018, this does not indicate that variation in ITN use is unimportant, as reported levels of use are often not reliable indicators of actual use. Overall, although improved tools will be needed in future, it is clear that substantive gains should still be achieved by more effective implementation of current policies on prevention and treatment, following WHO recommendations.

In addition to established methods, seasonal malaria chemoprevention (SMC) targeted to children under five years of age became a policy from 2014 onwards in several of the northern states where malaria prevalence has remained high, but only very limited implementation occurred in a few states before the 2018 survey. Community-based trials in diverse areas of West Africa where malaria is highly seasonal have demonstrated that the effect of intensively-delivered SMC on clinical incidence and parasite prevalence should be substantial ^24-26^. Although there are many challenges to optimal implementation in northern Nigeria where malaria is most seasonal, it is notable that SMC has been safely and effectively delivered to children up to the age of 10 years in trials in Senegal ^27^, so potential benefits of expanding the age coverage in Nigeria should be considered, while continuing efforts to establish effective delivery to younger children as currently intended.

Beyond disease-specific interventions, socio-economic development is needed to sustainably reduce malaria. In previous analysis of data from the 2015 MIS survey, many correlating variables were associated with malaria prevalence, including limited education of household heads and mothers, and unimproved housing ^12^. Although confounding socio-economic interactions prevent imputation of causality of individual determinants in such a survey, data from multiple studies elsewhere provide a consensus that house quality is a major determinant of malaria infection risk ^28 29^, as prevention of mosquito entry is a major means to reduce exposure. This is worthy of attention for research in Nigeria, as improvements in house design and construction are needed. Such improvements could bring the most substantive benefits to communities in rural areas, and it should be noted that they are separate to more general effects of urbanisation in locally reducing malaria ^30^. In addition, the aim for holistic implementation of malaria interventions across the country may not take account of the epidemiological differences among zones and states that may require more tailored intervention approaches. Aside from known ecological variation, and potential impacts of ongoing climate change that need to be determined, large areas of irrigation that support farming may extend breeding of vector mosquitoes and potentially require intervention all year round. Uncovering the local determinants of varying malaria transmission in each area may guide more appropriate interventions, and requires more formative research going forward.

Unscheduled disruptions also contribute to the overall challenges of malaria control. Sustained interventions in some states of the North East are have been difficult due to security challenges which impede access of commodities and health personnel for implementation and monitoring. More recently, disruption due to the Coronavirus pandemic is likely to cause reversals to the moderate reductions in malaria described here ^31^, and has already caused plans for the next national Malaria Indicator Survey that was scheduled for 2020 to be postponed. Given this setback, it is important to plan for more informative surveillance of malaria in Nigeria in future. For example, to supplement slide microscopy, molecular detection assays on DNA from dried blood spots would offer more highly sensitive and specific detection of infection, and would improve detection of rarer malaria parasite species that tend to occur as coinfections along with P. falciparum. The same samples could be used for monitoring of antimalarial drug resistance allele frequency changes, which will be a vital part of resistance management to direct policy on drugs to be used for antimalarial therapy ^32 33^ as well as other drugs for targeted prevention by SMC for young children ^27 34^ and intermittent preventive treatment for pregnant women ^34-36^ and potentially for infants. The benefits of such additional survey measures would become increasingly apparent over time, given the incremental value of repeated surveys that apply standardised laboratory methods as illustrated here.

## Abbreviations

(PR): Prevalence Ratio
(CI): Confidence Intervals
(PR_adj_): Mantel-Haenszel adjusted prevalence ratios
(WHO): World Health Organization
(DHS Program): Demographic and Health Surveys Program
(MIS): Malaria Indicator Survey
(NDHS): National Demographic and Health Survey
(ANDI): African Network for Drugs and Diagnostics Network
(ACT): artemisinin-based combination therapy
(ITN): insecticide treated net
(SMC): seasonal malaria chemoprevention.

## Contributors

WO and DJC planned and performed the analysis, and led the writing of the manuscript on behalf of all authors. OO contributed detailed input and suggestions on the interpretations and the presentation. WO supervised the laboratory microscopy and qualitative data checking. GN, PU, OO, NO, OA, FO, KM, SE, EN, CA, NE and MA gave input on the programmatic background to the surveys, and suggestions on interpretation of the findings in relation to malaria control.

## Data availability

All survey data were generated under the DHS Program (https://dhsprogram.com/), and may be accessed upon request to the UNICEF MICS team (http://mics.unicef.org/surveys).

## Statement on Ethics

Permission was granted by the UNICEF MICS team for this analysis, and the Ethics Committees of the participating institutions do not consider this to require separate approval, as it involves secondary analyses of anonymised data already in the public domain.

